# Comparing Three Evidence-Based Strategies to Reduce Cardiovascular Disease Burden: *An Individual-Based Cardiometabolic Policy Simulation*

**DOI:** 10.1101/2024.08.01.24311387

**Authors:** Sylvia Lutze, Steve Bachmeier, Alison Bowman, Nicole DeCleene, Hussain Jafari, Matthew Kappel, Caroline Kinuthia, Paulina Lindstedt, Megan Lindstrom, Rajan Mudambi, Christian Razo, Kjell Swedin, Abraham Flaxman, Gregory Roth

## Abstract

**• Background:** Understanding the real-world impact of clinical trials is important for informing health care policy. This is particularly true when trials are designed to show changes in surrogate endpoints such as changes in risk factors rather than events or mortality. We developed an agent-based microsimulation that estimates the population-level benefits in each US state for cardiometabolic health interventions shown to improve risk factors.

**• Methods:** We designed a large-scale, location-specific agent-based simulation model with a population of 51 million *in silico* individuals and estimated results for the years 2023 to 2040 in 30-day steps for each of the 50 states and District of Columbia. Input data reflected current cardiometabolic health in each state and the effects of interventions and risk factors on outcomes. We constructed three health policy intervention scenarios based on successful randomized controlled trials designed to improve cardiometabolic population health: improved access to fixed-dose combination (FDC) antihypertensive medication, a pharmacist-led intervention to increase adherence to statin and antihypertensives medications at the time they are initiated (Pharmacy), and a community-based lifestyle and behavior intervention designed to prevent diabetes (Community). Outcomes included myocardial infarction, ischemic and non-ischemic heart failure, and ischemic stroke events, deaths, and disability-adjusted life years (DALYs).

**• Results:** Our simulation included a representative population of the United States, accurate at the age, sex, and state level, with individual people simulated over 17 years. By the year 2040, the FDC intervention was estimated to have prevented 776,000 (95% UI 578,000– 956,000) CVD DALYs and 44,600 (95% UI 32,700–55,600) deaths annually. Reductions in ischemic heart disease deaths accounted for 76.5% of the total reductions in CVD deaths. The Pharmacist intervention prevented 170,000 (95% UI 129,000–208,000) CVD DALYs, and the Community intervention prevented 152,000 (95% UI 128,000–173,000) CVD DALYs.

**• Conclusions:** A fixed-dose combination of antihypertensives could prevent 1.2% of total CVD DALYs, with smaller benefits from adherence and lifestyle-focused programs and impact of interventions varying by state. The greatest reduction was in incident myocardial infarctions and ischemic heart disease deaths. Providing accurate population-level estimates at the state level can help local health policy decision-makers implement the most impactful interventions.

**Clinical Perspective:** **What is new?**

- Using person-level simulation, we have translated randomized trial results showing improvements in blood pressure, BMI, fasting plasma glucose, and LDL-C, and adherence to medication into real-world impact including forecasting cardiovascular disease events and deaths for the United States through the year 2040.
- A national, agent-based microsimulation for all 50 US states and DC allows us to assess how risk factor interventions will differentially affect demographic groups and locations.

**What are the clinical implications?**

- Broad adoption of fixed-dose combination medication for hypertension had the largest health benefit in all states.
- Interventions to improve adherence to medications or promote behavior change led to smaller reductions in disease burden.
- Direct comparison of the estimated real-world impact of clinical and community-based interventions can guide ongoing efforts to reduce the population burden of cardiovascular disease and resulting disparities.

## Introduction

Cardiovascular disease (CVD) is the leading cause of death in the United States and is responsible for 695,000 deaths annually,^1,2^ with up to a third of those estimated to be preventable.^3^ Large-scale health policy interventions such as the federally sponsored Million Hearts program have been rolled out to reduce cardiovascular disease burden, but impact has been limited.^4^ The strongest evidence supporting interventions to improve CVD comes from randomized clinical trials, which have shown benefits through medications, improved medication adherence, and behavioral changes. However, the real-world impact of these trials can be challenging to predict because they are most commonly powered for changes in surrogate endpoints such as risk factor levels, and their effect when implemented at scale in a state or health system can be unclear. Tools that estimate population-level impact from CVD interventions can provide policymakers with a realistic assessment of their choices and improve their ability to make necessary trade-off decisions between interventions.

To guide public health decision-making, we developed a person-level agent-based simulation for the entire United States. The simulation estimates the real-world impact of well-tested risk factor-modifying interventions on death, disability, and disease burden for state and national populations. Person-level agent-based simulation is a method borrowed from engineering and related fields that offers substantial advantages over traditional health models such as compartmental or decision tree simulations. For example, it can accurately represent complex, multi-step interventions common for cardiovascular diseases or even apply multiple interventions to be delivered over time, while also incorporating both the observed variation in individual response to therapies and uncertainty around that response.

In this analysis, we modeled 51 million individuals (“simulants”) to understand how interventions might change observed levels of risk factors in each US state, and how this change would affect cardiovascular events and deaths over the next two decades.

## Methods

### Overview

We constructed and ran an individual-based microsimulation for all 50 states and Washington, DC. Individual-based microsimulation is a computer modeling approach in which a cohort of 51 million *in silico* individuals was created with demographic and health characteristics, and those characteristics were changed sequentially in steps that represented the passage of time. Our simulation represented all adults over age 25 in each US state and ran in the simulated timeframe of 2023 through 2040. We utilized data from the Global Burden of Disease (GBD) 2019 study and other sources to match patterns of age, sex, metabolic risk factor exposure and burden, disease rate, mortality rate, and health care delivery in each US state.^5–8^ We designed the simulation to examine major metabolic risk factors: levels of systolic blood pressure (SBP), fasting plasma glucose (FPG), serum LDL cholesterol (LDL-C), and body mass index (BMI), and common cardiovascular diseases: ischemic heart disease (IHD), ischemic stroke, and both ischemic and non-ischemic heart failure. A high-level flow diagram can be seen in Figure 1, with detailed methods, an illustrative flow diagram (eFigure 1), and a schematic showing the relationship between risk factors and outcomes (eFigure 2) in the supplement. The study did not involve enrollment of human participants and used only existing population-level data.

**Figure 1:**
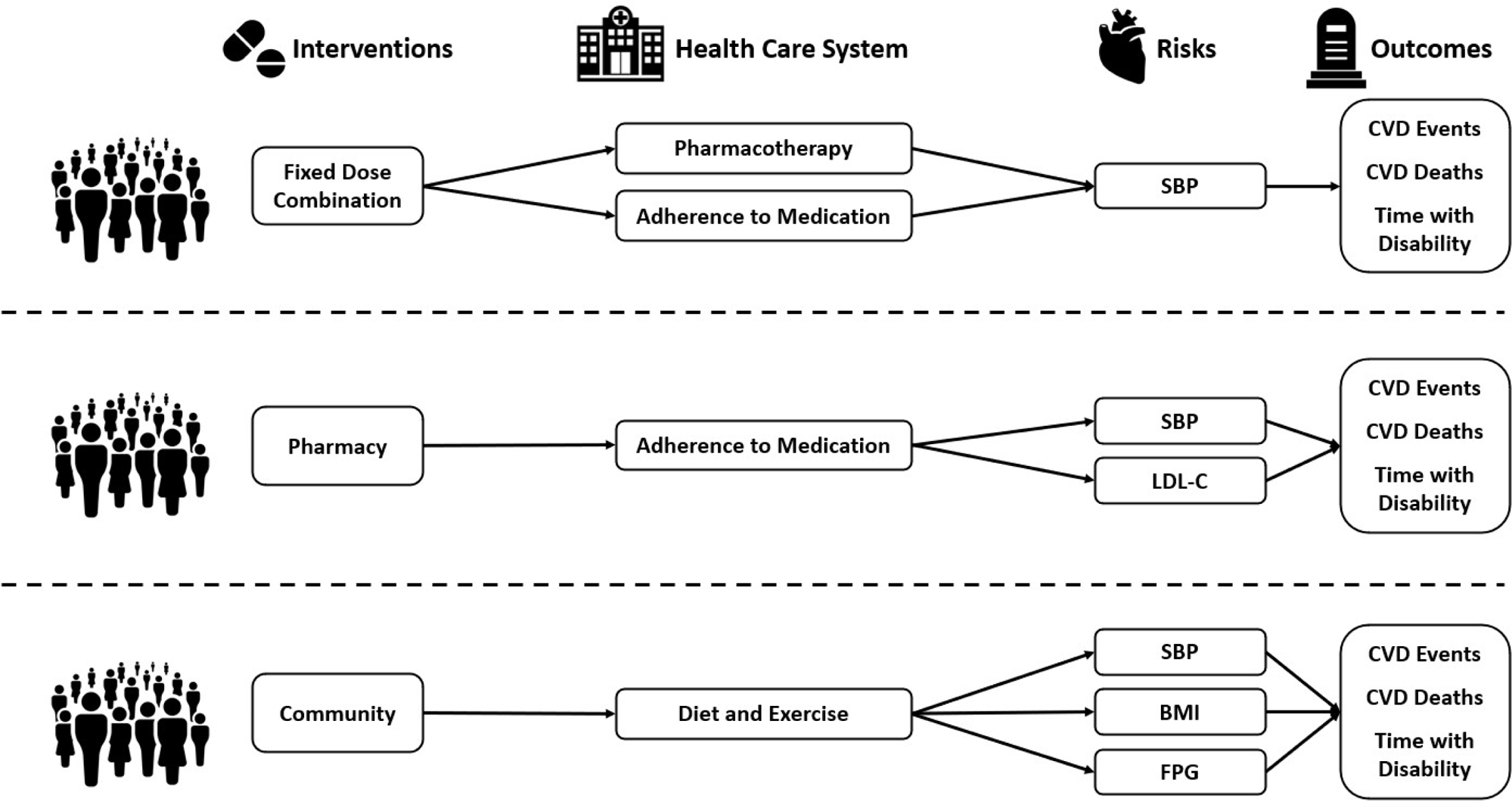
Flow of Simulants through the Model. *High-level overview of the flow of simulants, including the health care system, interventions, risks affected, and outcomes. More detailed information including data input values can be found in eFigure 1*.

### Input Data for the Simulation

Data for risk factor levels, disease incidence, and cause-specific mortality were obtained from the GBD study, along with existing published studies, including national and state-level population health surveys.^5–8^ The relationship between a simulant’s risk factor level and health outcomes, both initially and after treatment, were based on estimates of effect size from the GBD study’s meta-analyses of risk factor-outcome relationships.^9^ Further details on the GBD study’s estimation of relative risks can be found in the supplement and previous publication.^6^

At initialization of the simulation, each simulant received an age, sex, disease status, and location-specific level of systolic blood pressure (in mmHg), low-density lipoprotein cholesterol (in mmol/L), body mass index (in kg/m^2^), and fasting plasma glucose (in mmol/L) from GBD data. We calibrated health care visit rates to the GBD health care utilization study.^10^ To determine use of blood-pressure-lowering and cholesterol-lowering medications, we initialized simulants based on self-reported use in the National Health and Nutrition Examination Survey (NHANES) and Behavioral Risk Factor Surveillance System (BRFSS) data to reflect observed state variation in medication rates.^11,12^ Our model represented prescription and up-titration rates seen in real-world clinical practice, including AHA/ACC guideline-based practice as well as practices that diverge from that guidance.^13–15^ Our model also incorporated the effect of therapeutic inertia, the resistance to starting or increasing medication frequently observed in practice, using real-world rates reported in previously published studies.^16,17^ The biologic effect of pharmacotherapies on risk factors was based on large meta-analyses pooling the observed effect size across multiple studies.^18,19^ Real-world differences in patient adherence were included based on Medicare Part D data and a systematic review of the published literature.^20,21^ A full list of input values and data sources are included in supplement eTable 1.

### Individual-Based Simulation

We used the inputs above to create a simulation of risk factor exposures and disease rates through the year 2040. We modeled changes in the health of each simulant according to the following inputs: a) age- and sex-specific risk factor levels,^6^ disease and mortality rates,^5^ b) rates of primary care visits for risk factor screening,^10^ c) rates for the use of antihypertensive and lipid-lowering medications,^11^ d) rates of therapeutic inertia,^16,17^ and e) medication titration over time based on observed rates of follow-up and medication adjustment.^13–15,18–20^

We implemented this model using Vivarium, an open-source, Python-based simulation framework for our baseline health model.^22^ The baseline health model estimates a possible future trend in cardiovascular disease for each US state based on past CVD burden rates and expected changes in population, similar to other published forecasts.^23^ Published examples of models utilizing Vivarium can be found elsewhere.^24–26^ Simulated individuals, at each time step, are subject to the probability of a disease event, or health care event (appointment, medications prescribed, healthy individuals may experience CVD events, individuals experiencing CVD events may recover or die), which is modified by their other assigned attributes including age, sex, and cardiometabolic risk factor exposures. A high-level flow diagram can be seen in Figure 1, with a more detailed version as eFigure 1 in the supplement and medication effects on risk factors in eTables 2 and 3.

### Health Policy Intervention Scenarios

While many approaches have been tested over the past several decades to improve cardiovascular population health by modifying risk factors, for the purpose of this study we selected three interventions shown in high-quality randomized controlled trials to be effective for large populations. We intentionally selected interventions delivered in different locations (ambulatory clinics, pharmacies, communities) and targeting different combinations of risk factors to demonstrate the comparative effectiveness of substantially different interventions on a common set of cardiovascular outcomes. Also, comparison across these diverse settings for interventions has been prohibitively expensive and challenging to perform with prospective studies. We then modelled the impact of each intervention as if it were rolled out to an entire state’s population, to reflect the theoretical potential that could be achieved with maximum uptake of each intervention. We compared the baseline scenario described above to three counterfactual health policy intervention scenarios: a) broad adoption of a community-based lifestyle and behavior intervention similar to the US Diabetes Prevention Program delivered to all individuals who met the relevant entrance criteria of prediabetes (Community),^27,28^ b) broad adoption of a pharmacist-based intervention to increase medication adherence when a medication for high blood pressure or high cholesterol is first prescribed, delivered to all individuals who met the relevant entrance criteria (Pharmacy),^29^ or c) broad adoption of a fixed-dose combination antihypertensive intervention delivered to all individuals who met the relevant entrance criteria (FDC).^30–32^ Intervention effect reflects landmark trials that were designed with surrogate endpoints, cardiovascular risk factors, as the primary outcome. We modeled each intervention’s effect, via those surrogate endpoints, on myocardial infarction (MI), stroke, and heart failure event and mortality rates, as well as population-level burden as DALYs.

### Outcomes and Scenarios Analysis

For each scenario, we calculated rates of incident disease (myocardial infarction, stroke, heart failure), cause-specific death, all-cause death, years lived with disability (YLDs, years lived in less-than-ideal health), years of life lost (YLLs, years of life lost due to premature mortality), and disability-adjusted life years (DALYs, the sum of YLLs and YLDs) in each state, stratified by age and sex. We define CVD outcomes in this paper as the sum of ischemic stroke, ischemic heart disease, and heart failure. We compared intervention scenarios by calculating the number and percentage of cause-specific cardiac events, deaths, YLDs, YLLs, and DALYs averted compared to the baseline scenario. The simulation used identical simulants for each scenario who have the same probability of disease and death.^33^ State-level results were summed to national results based on state population. Results were averaged for annual values between 2025 and 2040 to capture the long-term impact of interventions once fully adopted.

### Validation and Data Processing

We ensured that model inputs remained stable within the complex dynamics of the simulation’s medications, CVD events, and mortality by comparing incidence, prevalence, and mortality rates for all diseases, risk factor distributions, and risk factor–attributable burden against GBD values.^5,6^ Additionally, we ensured health care visit rates, adherence rates, medication effects, and therapeutic inertia rates remained stable over time.^10,16–20^ Lastly, we ensured that verified that changes in adherence and risk factors seen in the simulation were the same as those seen in the RCT results.

In later years of the simulation, population sizes differ due to the interventions’ cumulative effects on lives saved. Capturing these increases in population are an important output of this type of simulation work. For the sake of clarity, counts data throughout this paper are scaled to the baseline population size. Further details are in the supplement.

### Statistical Analyses

We included parameter uncertainty in the microsimulation by running 10 replications with independent parameter values drawn from the posterior distribution of our input data, resulting in our modeling the life course of 51 million simulants. We used this to quantify uncertainty in our results by calculating the mean, range, and 95% uncertainty interval (95% UI) based on the different draw values.

## Results

### Estimated Reduction in Cardiovascular Disease DALYs

At the national level, the FDC intervention resulted in a 1.2% (95% UI 0.90–1.4%) reduction in CVD DALYs annually compared to the baseline scenario, while the Pharmacy intervention and the Community intervention resulted in 0.26% (95% UI 0.20–0.31%) and 0.23% (95% UI 0.20–0.26%) reductions, respectively. There was an average annual reduction between 2025 and 2040 of 776,000 (95% UI 578,000–956,000) DALYs in the FDC scenario, 170,000 (95% UI 129,000–208,000) in the Pharmacy intervention, and 152,000 (95% UI 128,000–173,000) in Community intervention nationwide (Table 1).

**Table 1:** Average Cardiovascular Disease Disability-Adjusted Life Years, Deaths, and Events Averted Annually in the United States between 2025 and 2040.

| Intervention | Sex | CVD DALYs Averted | CVD Deaths Averted | CVD Events Averted |  |  |
| --- | --- | --- | --- | --- | --- | --- |
|  |  |  |  | Myocardial Infarctions | Ischemic Strokes | Heart Failure Cases |
| FDC | Female | 362,000<br>(276,000 to 440,000) | 23,200<br>(17,100 to 28,700) | 26,300<br>(15,000 to 41,600) | 14,300<br>(7,600 to 19,600) | 11,900<br>(6,240 to 21,100) |
|  | Male | 415,000<br>(303,000 to 517,000) | 21,300<br>(15,600 to 27,000) | 31,300<br>(19,600 to 45,800) | 7,690<br>(4,880 to 10,600) | 9,690<br>(3,840 to 19,200) |
|  | **Total** | **776,000**<br>**(578,000 to 956,000)** | **44,600**<br>**(32,700 to 55,600)** | **57,700**<br>**(34,700 to 87,600)** | **22,100**<br>**(13,000 to 30,200)** | **21,600**<br>**(10,100 to 40,300)** |
| Pharmacy | Female | 76,600<br>(58,000 to 92,000) | 4,940<br>(3,690 to 6,080) | 8,240<br>(5,010 to 12,500) | 3,630<br>(2,340 to 5,040) | 802<br>(-268 to 2,110) |
|  | Male | 94,000<br>(69,000 to 116,000) | 4,590<br>(3,410 to 5,600) | 9,170<br>(5,670 to 12,500) | 1,890<br>(1,210 to 2,540) | 884<br>(58 to 2,130) |
|  | **Total** | **170,000**<br>**(129,000 to 208,000)** | **9,540**<br>**(7,100 to 11,600)** | **17,400**<br>**(10,700 to 25,100)** | **5,530**<br>**(3,700 to 7,520)** | **1,680**<br>**(216 to 4,240)** |
| Community | Female | 64,800<br>(52,800 to 74,500) | 3,860<br>(3,100 to 4,470) | 1,960<br>(1,000 to 3,300) | 719<br>(69 to 1,170) | 3,240<br>(2,450 to 4,010) |
|  | Male | 87,600<br>(70,800 to 102,000) | 4,400<br>(3,670 to 4,940) | 3,300<br>(2,160 to 4,840) | 495<br>(104 to 860) | 3,220<br>(2,500 to 4,230) |
|  | **Total** | **152,000**<br>**(128,000 to 173,000)** | **8,260**<br>**(6,860 to 9,420)** | **5,260**<br>**(3,360 to 8,150)** | **1,220**<br>**(173 to 1,990)** | **6,470**<br>**(5,330 to 8,250)** |

*Caption: Total cardiovascular disease (CVD) disability-adjusted life years (DALYs), CVD deaths, and CVD events* *averted annually between 2025 and 2040 under each alternative intervention scenario. Includes a 95% uncertainty* *interval based on variation between simulation runs.*

All states saw the highest impact from FDC interventions, with lower impacts from the Pharmacy intervention and Community intervention (Figure 2). The mean annual reduction in CVD DALYs from FDC varied between 1.5% and 0.97% across locations (Figure 2). For the Pharmacy intervention, the mean reduction in CVD DALYs varied between 0.32% and 0.19%, and for the Community intervention, it varied between 0.32% and 0.18%. Total DALYs averted by state can be found in eTable 7 in the supplement.

**Figure 2:**
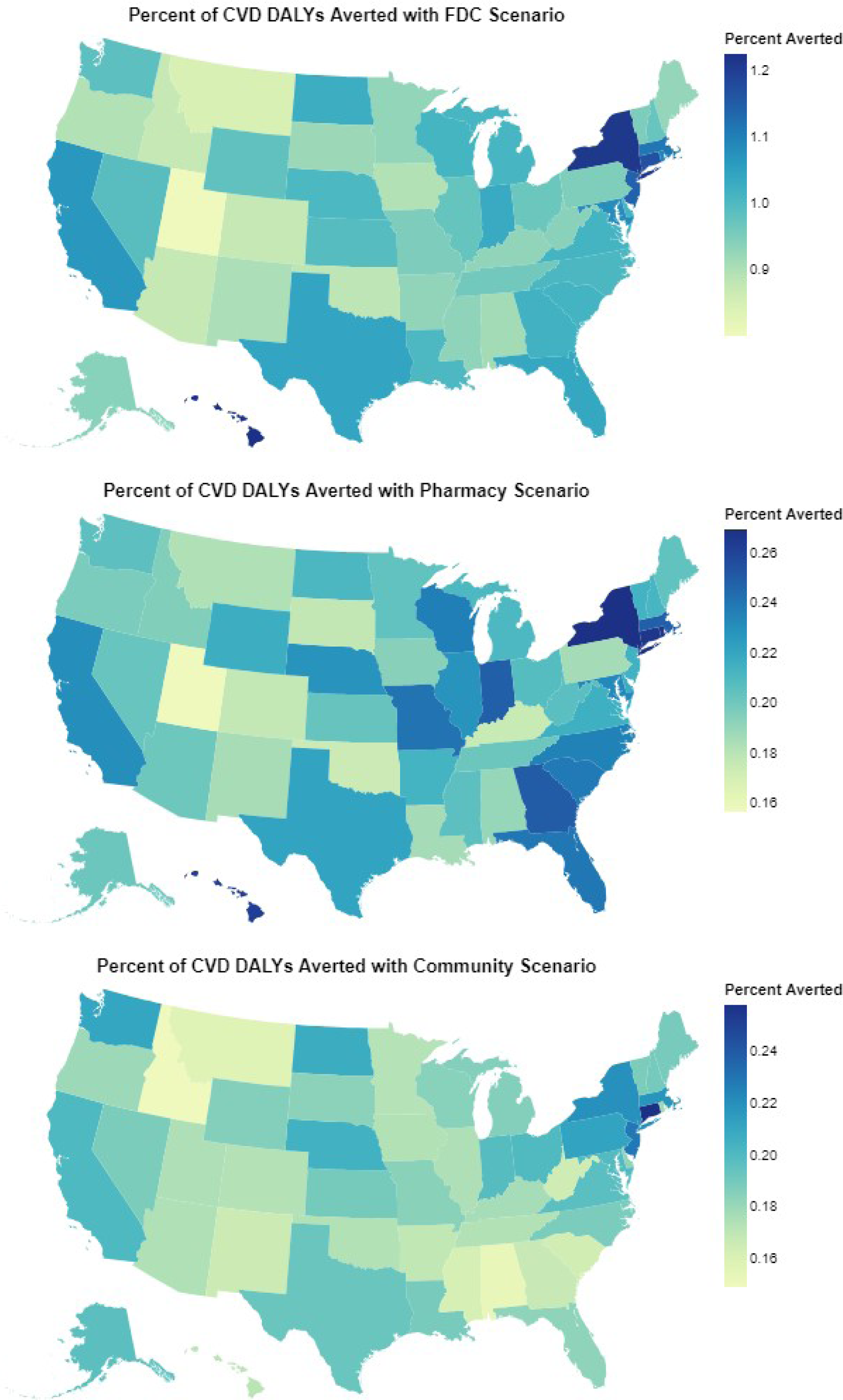
Map Showing CVD DALYs Averted from Each Intervention by State. *Percentage of cardiovascular disease (CVD) disability-adjusted life years (DALYs) averted under all intervention scenarios in each state*.

Interventions shifted the distribution of risk factors relative to the baseline scenario (Figure 3). We report California, Illinois, and Florida here as examples. Simulants who received the FDC intervention had an average 8.6 mmHg lower SBP in all states compared to the same simulants in the baseline scenario. Simulants who received the Pharmacy intervention had an SBP 0.38 to 0.42 mmHg lower and an LDL-C 0.33 to 0.34 mmol/L lower than baseline. Simulants who received the Community intervention had a 0.15 mmol/L lower FPG and 1.2 points lower BMI in all states.

**Figure 3:**
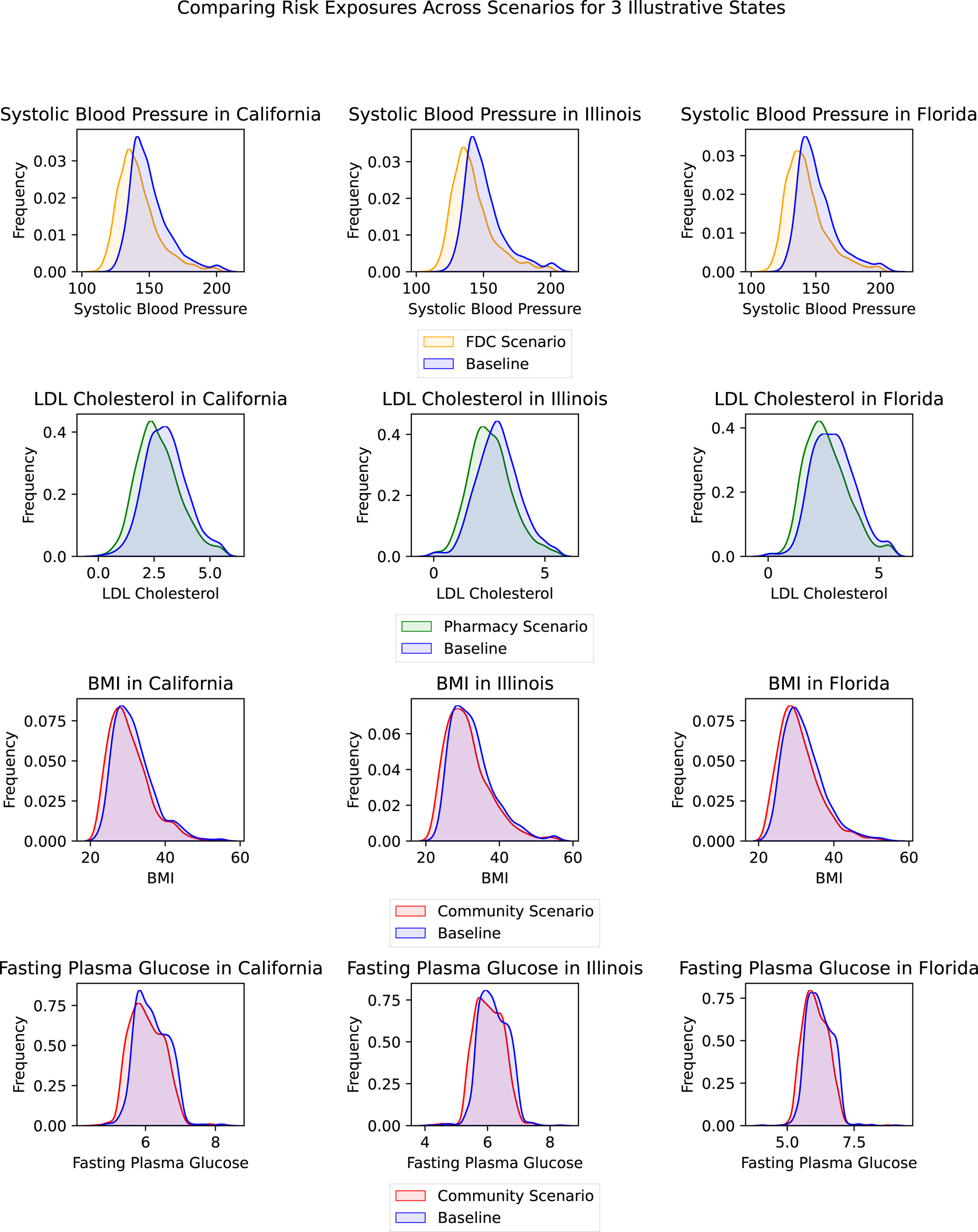
Risk Factor Distribution Comparison between Scenarios in Three States. *Change in risk factors between those who received each intervention and the same population in the baseline scenario. Shown for California, Illinois, and Florida. Only interventions that affect the risk factor are included in each graph. A version of* Figure 2 *for intention to treat can be found in eFigure 3 in the supplement*.

### Ischemic Heart Disease

The FDC intervention resulted in a 2.3% (95% UI 1.7–2.8%) annual reduction in IHD DALYs, compared to 0.55% (95% UI 0.39–0.66%) with the Pharmacy intervention and 0.37% (95% UI 0.32–0.42%) with the Community intervention. The FDC resulted in a 3.0% (95% UI 2.1–3.8%) annual reduction in IHD incidence and a 2.0% (95% UI 1.5–2.5%) decrease in IHD deaths. The Pharmacy intervention and the Community intervention, respectively, had a 0.79% (95% UI 0.54–1.0%) and 0.37% (95% UI 0.31–0.45%) reduction in IHD incidence, and a 0.48% (95% UI 0.33–0.59%) and 0.31% (95% UI 0.27–0.36%) reduction in IHD deaths.

The FDC intervention scenario averted 57,700 (95% UI 34,700–87,600) cases of myocardial infarction and 34,100 (95% UI 24,800–43,700) IHD deaths on average annually. The Pharmacy intervention and the Community intervention scenarios averted 17,400 (95% UI 10,700–25,100) and 5,260 (95% UI 3,360–8,160) cases of myocardial infarction and 8,190 (95% UI 5,490–10,600) and 5,330 (95% UI 4,420–6,180) IHD deaths. Total myocardial infarction count annually can be seen in Figure 4. State-level results are in eTables 5-7 in the supplement.

**Figure 4:**
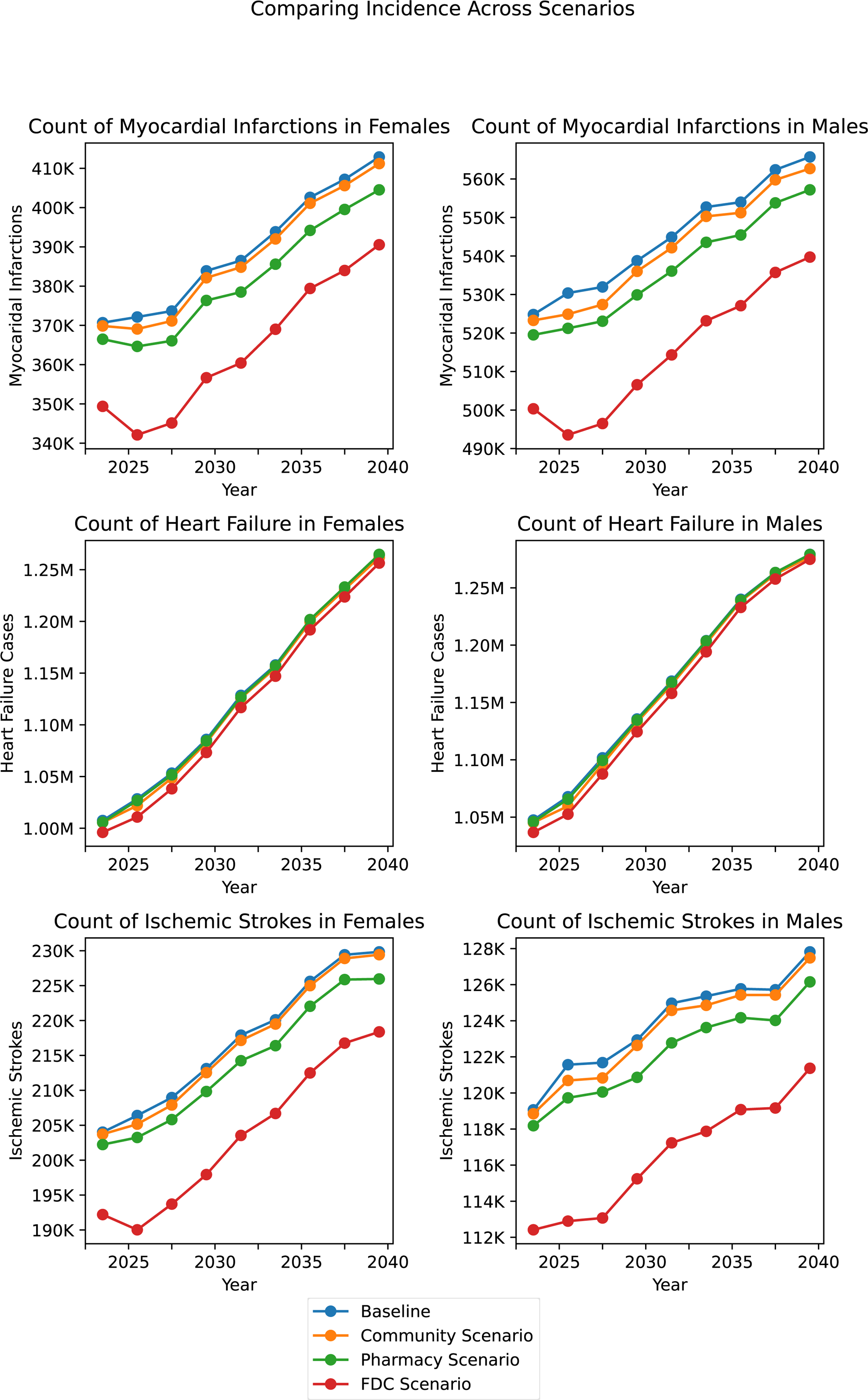
Line Graph for Count of Incident CVD Events between Scenarios Over Time. *Comparison of counts of total myocardial infarctions, heart failure diagnoses, and ischemic strokes per year for each intervention scenario*.

### Stroke

The FDC intervention resulted in a 2.1% (95% UI 0.89–2.7%) annual reduction in stroke DALYs. This led to 22,100 (95% UI 13,000–30,200) fewer strokes and 1,950 (95% UI 40–2,810) fewer stroke deaths nationwide. The Pharmacy intervention and Community interventions, respectively, resulted in a 0.58% (95% UI 0.36–0.85%) and 0.09% (95% UI −0.04–0.21%) reduction in stroke DALYs. This led to 5,530 (95% UI 3,700–7,520) and 1,220 (95% UI 176–1,990) fewer strokes and 664 (95% UI 253–1,110) and 63 (95% UI −76–229) fewer stroke deaths annually. Total stroke count annually can be seen in Figure 4. State-level results are in eTables 5-7 in the supplement.

### Heart Failure

There was a smaller reduction in heart failure cases compared to other CVD events (Figure 4). The FDC intervention resulted in a 1.0% (95% UI 0.64–1.6%) annual reduction in heart failure DALYs. This led to 21,600 (95% UI 10,100–40,300) fewer heart failure cases and 19,900 (95% UI 11,500–36,300) fewer heart failure deaths nationwide. The Pharmacy intervention and Community intervention resulted in a 0.13% (95% UI 0.08–0.21%) and 0.34% (95% UI 0.30–0.40%) reduction, respectively, in heart failure DALYs. This led to 1,680 (95% UI 216–4,240) and 6,470 (95% UI 5,330–8,250) fewer heart failure cases and 1,950 (95% UI 522–4030) and 6,170 (95% UI 5,280–7,740) fewer heart failure deaths annually. Total heart failure count annually can be seen in Figure 4. State-level results are in eTables 5-6 in the supplement.

### Age and Sex

Our input data were specific to 5-year age and sex groups, allowing us to assess how results might differ by population. Younger age groups saw much less benefit from interventions compared to ages 65 and above for all scenarios (Figure 5). The figure shows DALY counts averted, with the greatest reductions in 65–75- and 75–85-year-olds. Despite sex-specific risk factor and disease rate inputs, we did not see sex differences in relative impact or ordering of the interventions our results.

**Figure 5:**
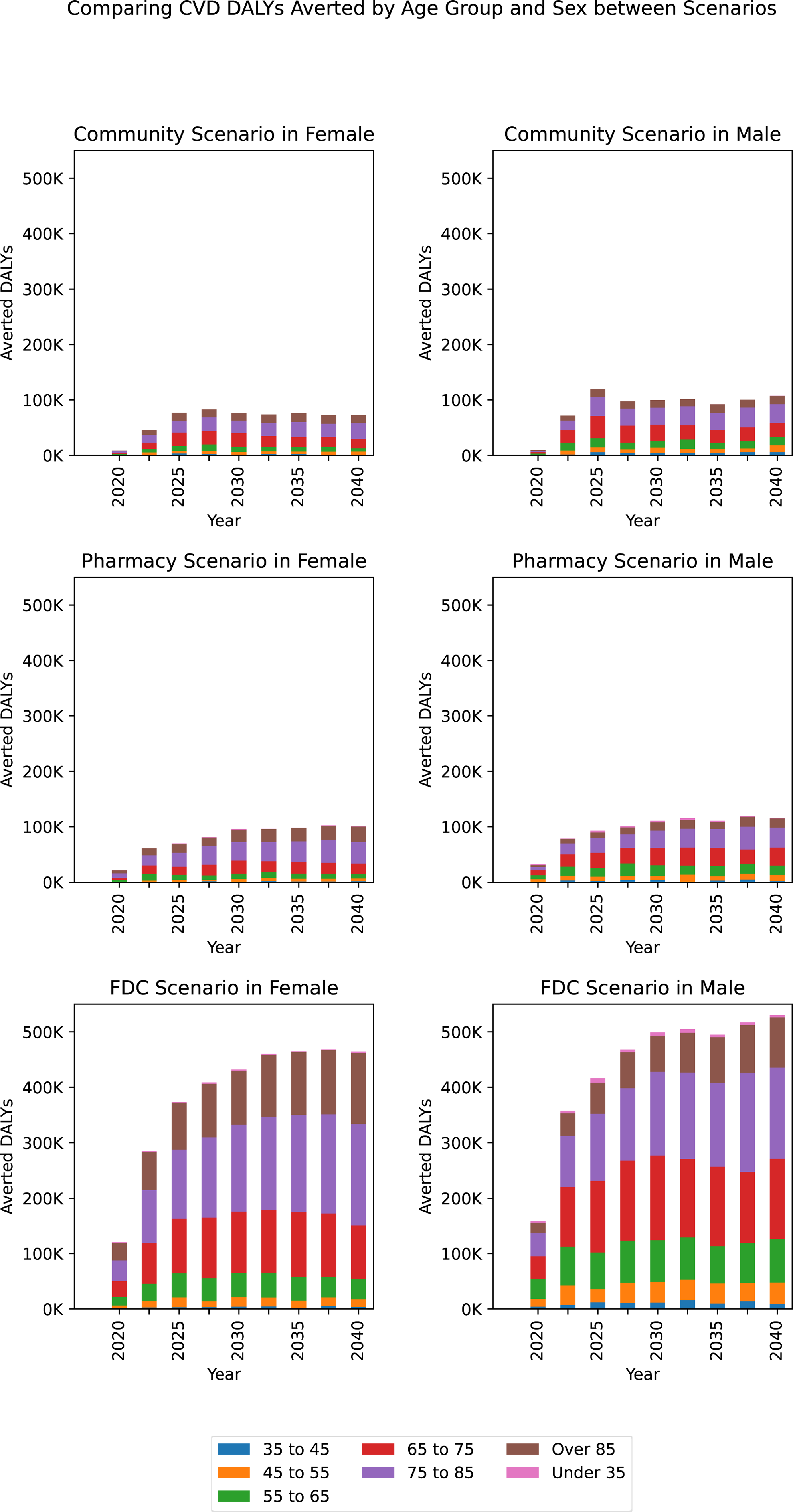
Stacked Bar Chart for CVD DALYs Averted between Scenarios by Age Group. *Comparison of CVD DALYs averted in each age group and for each intervention scenario: fixed-dose combination (FDC) antihypertensive, pharmacy intervention to increase medication adherence, and community-based National Diabetes Prevention Program (NDPP)*.

### Validation

Disease model inputs remained stable for the duration of the simulation, matching the empirically estimated disease rates from the Global Burden of Disease study used to initialize the first year of the simulation. Additionally, health care visit rates, adherence rates, medication effects, and therapeutic inertia rates remained stable over time and matched desired inputs. The stability of disease rates and health care activities is reassuring and shows that simulants continued to be exposed to a realistic process across many years of the simulation.

Our results were largely aligned with the results from the RCTs used to design the interventions. For example, the FDC intervention trials report medication adherence and change in SBP level, which were included directly in our simulation. Our results found an average decrease in SBP of 8.6 mmHg in all states analyzed 1 year into the simulation. The reference source FDC trials found similar results in blood pressure reduction.^31,32^

The intervention to increase medication adherence trial only reported change in adherence, which we directly implemented and modeled in our simulation. Similarly, for the Community intervention, we directly modeled changes in risk factors to match the initial and sustained drops seen in the 10-year follow-up of the NDPP. Further comparisons of simulated and RCT results can be found in eTable 4 in the supplement.

## Discussion

We estimate 4.0 million CVD events and 3.1 million CVD deaths, for a total of 67.8 million CVD DALYs in 2040 nationally. Our simulation suggests that substantial improvements in population health could be achieved with effective and broad delivery of proven interventions. Broad adoption of a fixed-dose combination antihypertensive regimen for the management of hypertension is estimated to prevent 101,300 cardiovascular events and 44,600 CVD deaths in the United States annually, and over 1.5 million events and 600,000 deaths by the year 2040. A pharmacy-based intervention to increase medication adherence is estimated to prevent 24,610 CVD events and 9,540 CVD deaths annually. A community-based intervention for lifestyle modification is estimated to prevent 12,950 CVD events and 8,260 CVD deaths annually. These latter two interventions achieved only about 25% of the reductions seen with the FDC intervention, suggesting that improvements in efficacy of pharmacotherapies would have a larger impact than broader adoption of the current federal program for lifestyle modification or interventions targeting improved medication adherence. Federal and state planning for population health goals should consider the relative benefits of interventions at the state and national level.

Our microsimulation emulated the intermediate outcomes found in 3 randomized clinical trials, and converted that impact into DALYs, events averted, and deaths averted within specific populations, including by sex, age, and state. These successful RCTs showed promising results, but rollout nationally was limited. Here, we show the significant public health benefit possible if these interventions are successfully scaled up. For example, successful implementation of the FDC intervention is estimated to achieve just under 40% of the federal government goal of preventing 1 million heart attacks and strokes over 5 years and would achieve the million reductions in approximately 13 years.^4^

Our health policy model is the first to provide separate forecasts for each US state. It also provides a comparison between multiple interventions. Prior policy models have been used to examine CVD health processes and outcomes.^34–36^ They have found similar-magnitude impacts for population-level interventions but only for single populations or locations.^37–39^ A state-level health policy model can help address decision-making at this important level of government. Recent years have seen state-level decisions on Medicaid funding and women’s health care that have serious implications for public health.^40,41^ Our model also can be updated with current disease rates with each update of the Global Burden of Disease study.

We observed large differences between the FDC scenario and the Pharmacy and Community scenarios. This impact gap reflects both the large effect size of the FDC intervention and the sizeable number of people in the total population eligible for this treatment, compared with the pharmacy- and community-based interventions. The FDC intervention results in larger reductions to SBP than have been shown with behavioral or lifestyle changes alone. Additionally, both hypertension and visits to health care providers are common in the US, meaning there were significantly more eligible people accessing the FDC intervention than the Community intervention, even accounting for the limiting effects of therapeutic inertia and nonadherence.^42^

The Pharmacy intervention was the second most effective. This intervention was designed to improve adherence rates to medications, resulting in a significant benefit for those simulants who began receiving medications. However, because the intervention only impacted individuals who would have been non-adherent, only 7% of antihypertensive medication users and 12% of cholesterol medication users experienced a benefit. The Community intervention program was limited, by design, to those who both have high BMI and are pre-diabetic, resulting in a smaller enrolled population size than the other interventions, approximately 2.8% of simulants compared to 9.6% in FDC intervention. Future community-based programs targeting behavior change should consider enrolling broader populations at risk.

Overall, heart failure cases saw less reduction than stroke or myocardial infarction (Figure 4). There are two reasons for this outcome. First, reductions in FPG and LDL-C have not been shown to reduce incident HF in longitudinal studies once changes in IHD are accounted for, and therefore this was not included in the simulation.^43–47^ The lack of observed effect may reflect limited sample size in large cohort studies, which suggests that any possible missed effect is small. Second, the attributable fraction of heart failure due to elevated SBP and BMI is much lower than in IHD and stroke. We estimated approximately 50% of IHD or stroke cases are caused by elevated BMI or SBP, but only about 20% of heart failure cases are caused by one of these risk factors.^6^

Our simulation utilizes data that are representative of the entire population in each state, which includes variation between states due to demographic differences. Analysis at the state level allows us to see how those demographic differences impact the relative impact from each intervention. The intervention effect was similar across all states, but the magnitude of disease burden averted varied widely. For example, Utah had the lowest rate of DALYs averted due to the FDC intervention, with 0.97%. Utah has one of the lowest average SBP levels of any state and one of the lowest rates of hypertensive medication use. Fewer simulants received the FDC intervention, and SBP is responsible for fewer CVD events in Utah, leading to a reduced impact.

Calibration of a simulation model to match real-world situations is essential to trusting the results of a computer-based simulation of population health. For example, our model reliably reproduced the expected results observed in RCTs. In the FDC intervention, our model found SBP reductions within the confidence interval of 2 of the 3 trials for combination therapies reviewed.^31,32^ This is especially important as the change in SBP was based on medication prescription and dynamics within the model, not based on a direct effect of the intervention. This means that the complex health care system designed to emulate real-world medication dynamics was successful in capturing sufficient detail.

### Limitations

Given the scale of data inputs required for a simulation of this kind, some necessary input data were limited in level of available detail or geographic representation. This was most apparent for inputs used to model aspects of the health care system, such as therapeutic inertia or medication adherence. Although we have age/sex/state-specific values for incidence, mortality rates, risk factors, and medication rates, we assumed that the rates of adherence and therapeutic inertia did not vary by age, sex, or state. For these limited components where we did not reflect possible variation in input data, we might not fully capture expected variation in outcomes by sex, age, or state. We would not expect this to bias our results at the aggregate, but our estimates could diverge from real-world when input is limited in this way. We do note that there are very few population-level data available on medication adherence or therapeutic inertia, and we do not know if age, sex, or location has a substantial impact on these aspects of health care delivery. Also, our choice to estimate intervention impact using RCTs may lead to overestimation of their impact due to differences between efficacy of an intervention among a highly selected population enrolled in a trial and effectiveness in the general population. We accepted this as a potential source of bias in order to inform the simulation with the most reliable and robust estimates of each intervention’s effect, based on estimates that due to randomization were unaffected by either observed or unobserved confounding.^48^

## Conclusion

This simulation quantifies the potential public health benefit if effective interventions were to be scaled up successfully at the state and national level. Broad adoption of a fixed-dose combination antihypertensive regimen for the management of hypertension was estimated to prevent 101,300 CVD events and 44,600 CVD deaths in the United States annually, and over 1.5 million events and 600,000 deaths by the year 2040. This simulation suggests that substantial improvements in population health are achievable with effective and broad delivery of proven interventions.

## Data Availability

The dataset generated for use during the current study is available in the ihmeuw/vivarium_nih_us_cvd repository, the link for the simulation data is: https://zenodo.org/doi/10.5281/zenodo.10671674

https://github.com/ihmeuw/vivarium_nih_us_cvd

https://zenodo.org/doi/10.5281/zenodo.10671674

## References

1. Multiple Cause of Death Data on CDC WONDER. Accessed November 22, 2023. https://wonder.cdc.gov/mcd.html

2. Heart Disease and Stroke Statistics—2023 Update: A Report From the American Heart Association | Circulation. Accessed November 22, 2023. https://www.ahajournals.org/doi/10.1161/CIR.0000000000001123

3. Potentially Preventable Deaths from the Five Leading Causes of Death — United States, 2008–2010. Accessed November 22, 2023. https://www.cdc.gov/mmwr/preview/mmwrhtml/mm6317a1.htm

4. CDC. Million Hearts®. Centers for Disease Control and Prevention. Published May 5, 2023. Accessed November 14, 2023. https://millionhearts.hhs.gov/index.html

5. Vos T, Lim SS, Abbafati C, et al. Global burden of 369 diseases and injuries in 204 countries and territories, 1990–2019: a systematic analysis for the Global Burden of Disease Study 2019. The Lancet. 2020;396(10258):1204–1222. doi:10.1016/S0140-6736(20)30925-9

6. Murray CJL, Aravkin AY, Zheng P, et al. Global burden of 87 risk factors in 204 countries and territories, 1990–2019: a systematic analysis for the Global Burden of Disease Study 2019. The Lancet. 2020;396(10258):1223–1249. doi:10.1016/S0140-6736(20)30752-2

7. Kenchaiah S, Sesso HD, Gaziano JM. Body Mass Index and Vigorous Physical Activity and the Risk of Heart Failure Among Men. Circulation. 2009;119(1):44–52. doi:10.1161/CIRCULATIONAHA.108.807289

8. Zhang Y, Vittinghoff E, Pletcher MJ, et al. Associations of Blood Pressure and Cholesterol Levels During Young Adulthood With Later Cardiovascular Events. J Am Coll Cardiol. 2019;74(3):330–341. doi:10.1016/j.jacc.2019.03.529

9. Zheng P, Afshin A, Biryukov S, et al. The Burden of Proof studies: assessing the evidence of risk. Nat Med. 2022;28(10):2038–2044. doi:10.1038/s41591-022-01973-2

10. Moses MW, Pedroza P, Baral R, et al. Funding and services needed to achieve universal health coverage: applications of global, regional, and national estimates of utilisation of outpatient visits and inpatient admissions from 1990 to 2016, and unit costs from 1995 to 2016. Lancet Public Health. 2019;4(1):e49–e73. doi:10.1016/S2468-2667(18)30213-5

11. Centers for Disease Control and Prevention (CDC) National Center for Health Statistics (NCHS). National Health and Nutrition Examination Survey Data. https://wwwn.cdc.gov/nchs/nhanes/continuousnhanes/default.aspx?BeginYear=2017

12. CDC - BRFSS. Published January 9, 2024. Accessed January 25, 2024. https://www.cdc.gov/brfss/index.html

13. Arnett DK, Blumenthal RS, Albert MA, et al. 2019 ACC/AHA Guideline on the Primary Prevention of Cardiovascular Disease: Executive Summary: A Report of the American College of Cardiology/American Heart Association Task Force on Clinical Practice Guidelines. Circulation. 2019;140(11). doi:10.1161/CIR.0000000000000677

14. Byrd JB, Zeng C, Tavel HM, et al. Combination therapy as initial treatment for newly diagnosed hypertension. Am Heart J. 2011;162(2):340–346. doi:10.1016/j.ahj.2011.05.010

15. Nguyen V, deGoma EM, Hossain E, Jacoby DS. Updated cholesterol guidelines and intensity of statin therapy. J Clin Lipidol. 2015;9(3):357–359. doi:10.1016/j.jacl.2014.12.009

16. Turchin A, Goldberg SI, Shubina M, Einbinder JS, Conlin PR. Encounter Frequency and Blood Pressure in Hypertensive Patients with Diabetes. Hypertension. 2010;56(1):68–74. doi:10.1161/HYPERTENSIONAHA.109.148791

17. Goldberg KC, Melnyk SD, Simel DL. Overcoming inertia: improvement in achieving target low-density lipoprotein cholesterol. Am J Manag Care. 2007;13(9):530–534.

18. Law MR, Morris JK, Wald NJ. Use of blood pressure lowering drugs in the prevention of cardiovascular disease: meta-analysis of 147 randomised trials in the context of expectations from prospective epidemiological studies. BMJ. 2009;338:b1665. doi:10.1136/bmj.b1665

19. Descamps O, Tomassini JE, Lin J, et al. Variability of the LDL-C lowering response to ezetimibe and ezetimibe + statin therapy in hypercholesterolemic patients. Atherosclerosis. 2015;240(2):482–489. doi:10.1016/j.atherosclerosis.2015.03.004

20. Cheen MHH, Tan YZ, Oh LF, Wee HL, Thumboo J. Prevalence of and factors associated with primary medication non-adherence in chronic disease: A systematic review and meta-analysis. Int J Clin Pract. 2019;73(6):e13350. doi:10.1111/ijcp.13350

21. Part C and D Performance Data | CMS. Accessed January 30, 2024. https://www.cms.gov/medicare/health-drug-plans/part-c-d-performance-data

22. Vivarium. https://vivarium.readthedocs.io/en/latest/

23. Heidenreich PA, Albert NM, Allen LA, et al. Forecasting the impact of heart failure in the United States: a policy statement from the American Heart Association. Circ Heart Fail. 2013;6(3):606–619. doi:10.1161/HHF.0b013e318291329a

24. Haddock B, Pletcher A, Blair-Stahn N, et al. Simulated data for census-scale entity resolution research without privacy restrictions: a large-scale dataset generated by individual-based modeling. Gates Open Res. 2024;8:36. doi:10.12688/gatesopenres.15418.1

25. Kannan A, Tsoi D, Xie Y, Horst C, Collins J, Flaxman A. Cost-effectiveness of Vitamin A supplementation among children in three sub-Saharan African countries: An individual-based simulation model using estimates from Global Burden of Disease 2019. Horton S, ed. PLOS ONE. 2022;17(4):e0266495. doi:10.1371/journal.pone.0266495

26. Young N, Bowman A, Swedin K, et al. Cost-effectiveness of antenatal multiple micronutrients and balanced energy protein supplementation compared to iron and folic acid supplementation in India, Pakistan, Mali, and Tanzania: A dynamic microsimulation study. PLOS Med. 2022;19(2):e1003902. doi:10.1371/journal.pmed.1003902

27. Diabetes Prevention Program Outcomes Study Research Group, Orchard TJ, Temprosa M, et al. Long-term effects of the Diabetes Prevention Program interventions on cardiovascular risk factors: a report from the DPP Outcomes Study. Diabet Med J Br Diabet Assoc. 2013;30(1):46–55. doi:10.1111/j.1464-5491.2012.03750.x

28. National Diabetes Prevention Program | Diabetes | CDC. Published December 27, 2022. Accessed February 1, 2023. https://www.cdc.gov/diabetes/prevention/index.html

29. Derose SF, Green K, Marrett E, et al. Automated Outreach to Increase Primary Adherence to Cholesterol-Lowering Medications. JAMA Intern Med. 2013;173(1):38–43. doi:10.1001/2013.jamainternmed.717

30. Thom S, Poulter N, Field J, et al. Effects of a Fixed-Dose Combination Strategy on Adherence and Risk Factors in Patients With or at High Risk of CVD: The UMPIRE Randomized Clinical Trial. JAMA. 2013;310(9):918–929. doi:10.1001/jama.2013.277064

31. Muñoz D, Uzoije P, Reynolds C, et al. Polypill for Cardiovascular Disease Prevention in an Underserved Population. N Engl J Med. 2019;381(12):1114–1123. doi:10.1056/NEJMoa1815359

32. Webster R, Salam A, de Silva HA, et al. Fixed Low-Dose Triple Combination Antihypertensive Medication vs Usual Care for Blood Pressure Control in Patients With Mild to Moderate Hypertension in Sri Lanka: A Randomized Clinical Trial. JAMA. 2018;320(6):566–579. doi:10.1001/jama.2018.10359

33. Flaxman AD, Deason AW, Dolgert AJ, et al. Untangling uncertainty with common random numbers: a simulation study. In: Proceedings of the Summer Simulation Multi-Conference. SummerSim ‘17. Society for Computer Simulation International; 2017:1–12.

34. Weinstein MC, Coxson PG, Williams LW, Pass TM, Stason WB, Goldman L. Forecasting coronary heart disease incidence, mortality, and cost: the Coronary Heart Disease Policy Model. Am J Public Health. 1987;77(11):1417–1426. doi:10.2105/AJPH.77.11.1417

35. Pandya A, Sy S, Cho S, Alam S, Weinstein MC, Gaziano TA. Validation of a Cardiovascular Disease Policy Micro-simulation Model Using Both Survival and Receiver Operating Characteristic Curves. Med Decis Mak Int J Soc Med Decis Mak. 2017;37(7):802–814. doi:10.1177/0272989X17706081

36. Hirsch G, Homer J, Evans E, Zielinski A. A System Dynamics Model for Planning Cardiovascular Disease Interventions. Am J Public Health. 2010;100(4):616–622. doi:10.2105/AJPH.2009.159434

37. Kazi DS, Wei PC, Penko J, et al. Scaling Up Pharmacist-Led Blood Pressure Control Programs in Black Barbershops: Projected Population Health Impact and Value. Circulation. 2021;143(24):2406–2408. doi:10.1161/CIRCULATIONAHA.120.051782

38. Bibbins-Domingo K, Chertow GM, Coxson PG, et al. Projected effect of dietary salt reductions on future cardiovascular disease. N Engl J Med. 2010;362(7):590–599. doi:10.1056/NEJMoa0907355

39. Bellows BK, Ruiz-Negrón N, Bibbins-Domingo K, et al. Clinic-based strategies to reach United States Million Hearts 2022 blood pressure control goals: a simulation study. Circ Cardiovasc Qual Outcomes. 2019;12(6):e005624. doi:10.1161/CIRCOUTCOMES.118.005624

40. Bossick AS, Brown J, Hanna A, Parrish C, Williams EC, Katon JG. Impact of State-Level Reproductive Health Legislation on Access to and Use of Reproductive Health Services and Reproductive Health Outcomes: A Systematic Scoping Review in the Affordable Care Act Era. Womens Health Issues. 2021;31(2):114–121. doi:10.1016/j.whi.2020.11.005

41. Woolf SH. The Growing Influence of State Governments on Population Health in the United States. JAMA. 2022;327(14):1331–1332. doi:10.1001/jama.2022.3785

42. CDC. Hypertension Prevalence in the U.S. | Million Hearts®. Centers for Disease Control and Prevention. Published May 12, 2023. Accessed November 14, 2023. https://millionhearts.hhs.gov/data-reports/hypertension-prevalence.html

43. Lee MMY, Sattar N, McMurray JJV, Packard CJ. Statins in the Prevention and Treatment of Heart Failure: a Review of the Evidence. Curr Atheroscler Rep. 2019;21(10):41. doi:10.1007/s11883-019-0800-z

44. Agarwala A, Pokharel Y, Saeed A, et al. The association of lipoprotein(a) with incident heart failure hospitalization: Atherosclerosis Risk in Communities study. Atherosclerosis. 2017;262:131–137. doi:10.1016/j.atherosclerosis.2017.05.014

45. Bell DSH, Goncalves E. Heart failure in the patient with diabetes: Epidemiology, aetiology, prognosis, therapy and the effect of glucose-lowering medications. Diabetes Obes Metab. 2019;21(6):1277–1290. doi:10.1111/dom.13652

46. Nichols GA, Koro CE, Kolatkar NS. The incidence of heart failure among nondiabetic patients with and without impaired fasting glucose. J Diabetes Complications. 2009;23(3):224–228. doi:10.1016/j.jdiacomp.2007.10.001

47. Deedwania P, Patel K, Fonarow GC, et al. Prediabetes is not an independent risk factor for incident heart failure, other cardiovascular events or mortality in older adults: Findings from a population-based cohort study. Int J Cardiol. 2013;168(4):3616–3622. doi:10.1016/j.ijcard.2013.05.038

48. Nordon C, Karcher H, Groenwold RHH, et al. The “Efficacy-Effectiveness Gap”: Historical Background and Current Conceptualization. Value Health. 2016;19(1):75–81. doi:10.1016/j.jval.2015.09.2938

